# Reduced mortality among COVID-19 ICU patients after treatment with HemoClear convalescent plasma in Suriname

**DOI:** 10.1101/2022.12.10.22283287

**Authors:** R. Bihariesingh-Sanchit, R. Bansie, N. Ramdhani, R. Mangroo, D. Bustamente, E. Diaz, C. Fung A Foek, I. Thakoer, S. Vreden, Z. Choudhry, A.B. van ‘t Wout, D.A. Diavatopoulos, A.P. Nierich

## Abstract

Convalescent plasma is a promising therapy for coronavirus disease 2019 (COVID-19), but its efficacy in intensive care unit (ICU) patients in a low and middle income country setting such as Suriname is unknown. Bedside plasma separation using the HemoClear device made convalescent plasma therapy accessible as treatment option in Suriname. Two hundred patients with severe SARS-CoV-2 infection requiring intensive care were recruited. Fifty eight patients (29%) received COVID-19 convalescent plasma (CCP) treatment in addition to standard of care (SOC). The CCP treatment and SOC groups were matched by age, sex, and disease severity scores. Mortality in the CCP treatment group was significantly lower than in the SOC group (21% versus 39%; Fisher’s exact *P* = 0.0133). Multivariate analysis using ICU days showed that CCP treatment reduced mortality (hazard ratio [HR], 0.35; 95% CI, 0.18–0.66; *P* = 0.001), while complication of acute renal failure (creatinine levels >110 mol/L; HR, 4.45; 95%CI, 2.54-7.80; *P* < 0.0001) was independently associated with death. Decrease in chest X-ray score in the CCP treatment group (median -3 points, IQR -4 to -1) was significantly greater than in the SOC group (median -1 point, IQR -3 to 1, Mann Whitney *P* = 0.0004). Improvement in PaO2/FiOs ratio was also significantly greater in the CCP treatment group (median 83, IQR 8 to 140) than in the SOC group (median 35, IQR -3 to 92, Mann Whitney *P* = 0.0234). Further research is needed for HemoClear-produced CCP as therapy in SARS-CoV-2 infections together with adequately powered, randomized controlled trials.

**Importance:** This study compares mortality and other endpoints between intensive care unit (ICU) COVID-19 patients treated with convalescent plasma plus standard of care (CCP), and a control group of patients hospitalized in the same medical ICU facility treated with standard of care alone (SOC) in a low and middle income country (LMIC) setting using bedside donor whole blood separation by gravity (HemoClear) to produce the CCP. It demonstrates a significant 65% survival improvement in HemoClear-produced CCP recipients (HR 0.35; 95% CI, 0.19–0.66; P = 0.001). Although this is an exploratory study, it clearly shows the benefit of using the HemoClear-produced CCP in ICU patients in the Suriname LMIC setting. Additional studies can further substantiate our findings and their applicability to both LMICs and high income countries and the use of CCP as a prepared readiness method to combat new viral pandemics.

## Introduction

As of 28 November 2022, the COVID-19 pandemic has resulted in 637 million confirmed cases and 6.6 million deaths globally (WHO, https://covid19.who.int/) since the discovery of the Severe Acute Respiratory Syndrome Coronavirus 2 (SARS-CoV-2) in December 2019 (1, 2). Clinical manifestation ranges from mild upper respiratory tract illness to a diffuse viral pneumonia causing acute respiratory failure, multiorgan dysfunction and death. The absence of effective therapies prompted use of COVID-19 convalescent plasma (CCP) because of the historical efficacy of convalescent plasma treatment in human respiratory viral infections (3–5). Although early CCP treatment of hospitalized patients with COVID-19 reduced mortality in matched-control studies (6–9), randomized clinical trials have yielded mixed results, reducing mortality in one study (10), but not others (11–15)(16) despite showing signals of efficacy in subgroups.

With a population just over 500,000 inhabitants and neighboring Brazil where high COVID-19 disease incidence was reported, Suriname was confronted with a second COVID-19 wave at the end of May 2020. Given limited treatment options, we initiated a clinical trial to evaluate clinical efficacy of CCP treatment in patients admitted to the Intensive Care Unit (ICU) with severe or life-threatening COVID-19 in Suriname (SurCovid trial)(17). Because of the low resource setting and lack of conventional plasmapheresis machines, the HemoClear gravity-driven blood filter was used for CCP production (18). Here, we present the results of CCP treatment on the primary outcome of mortality in COVID-19 ICU patients in Suriname during the second wave during the COVID-19 pandemic.

## Methods

This work is reported in adherence to the preferred Strengthening the Reporting of Observational studies in Epidemiology (STROBE) guidelines. This prospective cohort study was performed at the Intensive Care Unit (ICU) of Academic Hospital Paramaribo and the Wanica Regional Hospital, Suriname, from June 2020 to October 2021 (ISRCTN49832318, https://doi.org/10.1186/ISRCTN49832318). Two hundred patients were included.

### Trial design and oversight

In this prospective cohort study, we compared CCP treatment with standard of care versus standard of care alone in patients with severe COVID-19 admitted to the ICU. After referral to the ICU, patients were treated with additional respiratory and circulatory support. After being found eligible for additional CCP treatment, a selection to either standard of care treatment including dexamethasone or standard treatment combined with CCP was started. The study flow chart in (**Figure 1**) illustrates the study enrolment and design.

**Figure 1.**
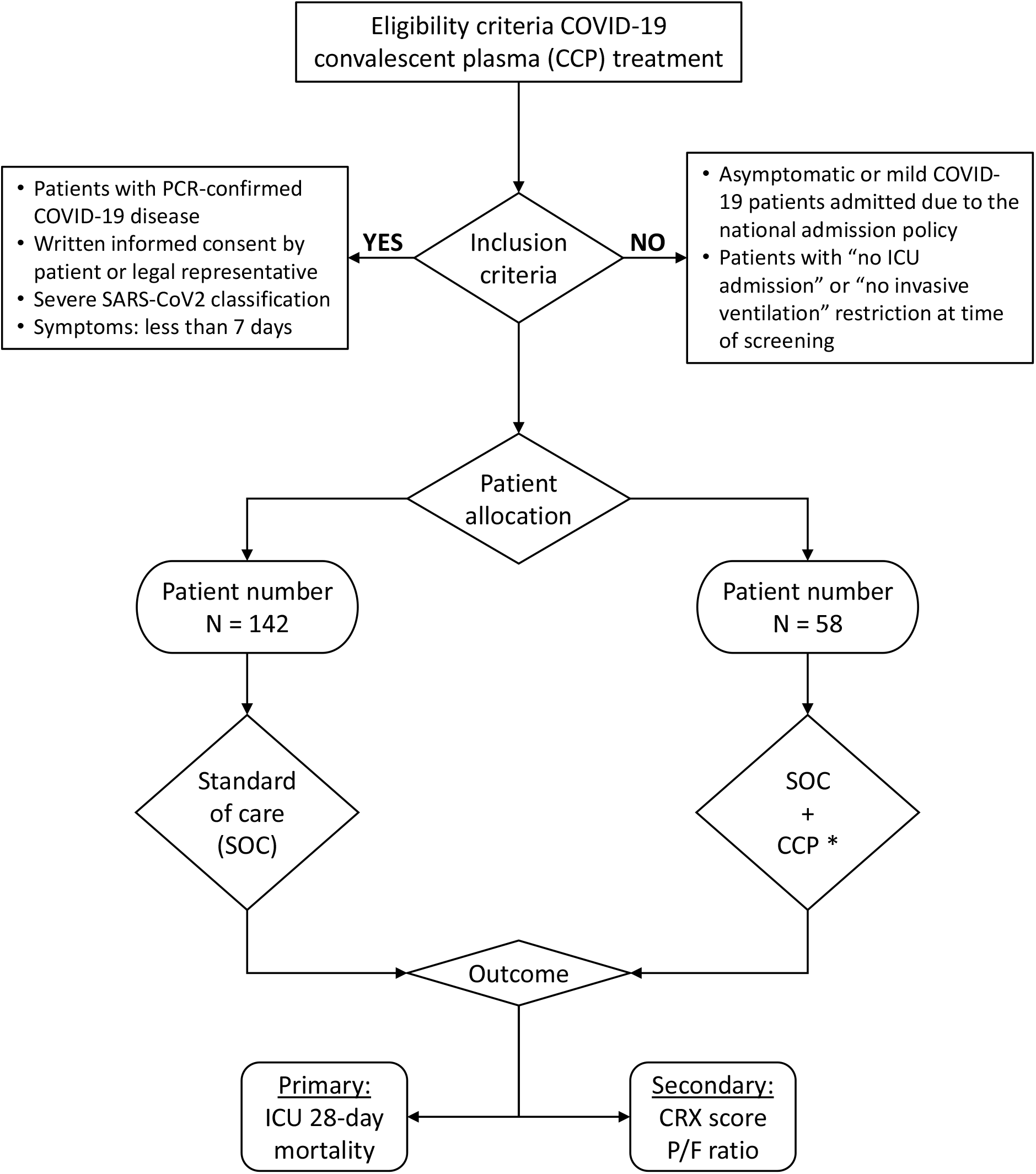
Patient selection flowchart with study enrolment and design. SOC: Oral or intravenous dexamethasone once daily. CCP: 2 units of COVID-19 convalescent plasma. Each unit infused over a period of at least 30 minutes. Close observation during the entire time of infusion. Adverse event medication (anaphylaxis, antihistamines, corticosteroids and epinephrine) within reach.

Ethical approval was granted by the Suriname Ministry of Health’s Ethics Review Board (registration number: IGAP02-482020; ISRCTN18304314). The data used to support the findings of this study are available from the corresponding author upon request.

### Patient population

Consenting adult patients (>18 years) with severe COVID-19 were enrolled in the trial in the period from June 2020 until October 2021. The eligibility criteria included written informed consent given by the patient or next of kin, a PCR confirmed diagnosis of COVID-19, and admittance to the ICU due to progressive respiratory failure ranging between severe and life-threatening acute respiratory distress syndrome (ARDS) based on the Berlin classification (19). For the interventional CCP group, all patients admitted at the ICU who met the inclusion criteria were approached and 58 patients who consented to CCP infusion plus standard of care therapy were recruited to this arm. For the control group with standard of care therapy alone (SOC treatment group), patients were included who did not consent to CCP infusion or when there was no CCP available (n=142). To account for major confounding factors, the following variables were used: age, gender, co-morbidities including the history of diabetes mellitus, hypertension, drugs, symptoms and signs (**Table 1**). In addition, both the SOC and CCP treatment groups received the same standard of care concurrent treatment, which included administration of oral or intravenous dexamethasone once daily, for up to 10 days. CCP treatment group patients were infused with two units of 220 mL CCP. For plasma selection, ABO compatibility was considered, regardless of Rhesus factor status. CCP recipients were monitored for serious adverse effects (SAEs) of CCP transfusion, including anaphylaxis.

**Table 1:**
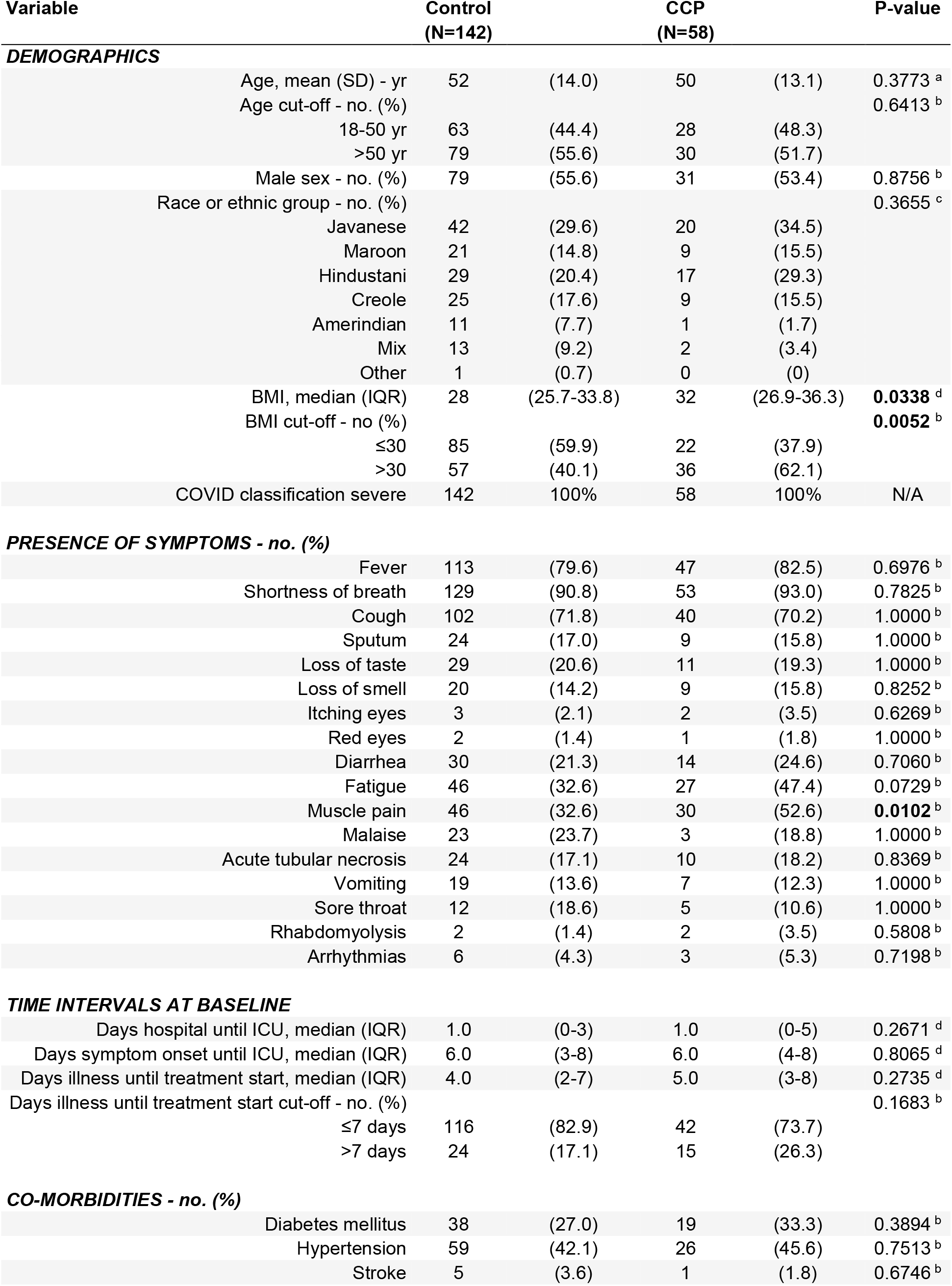
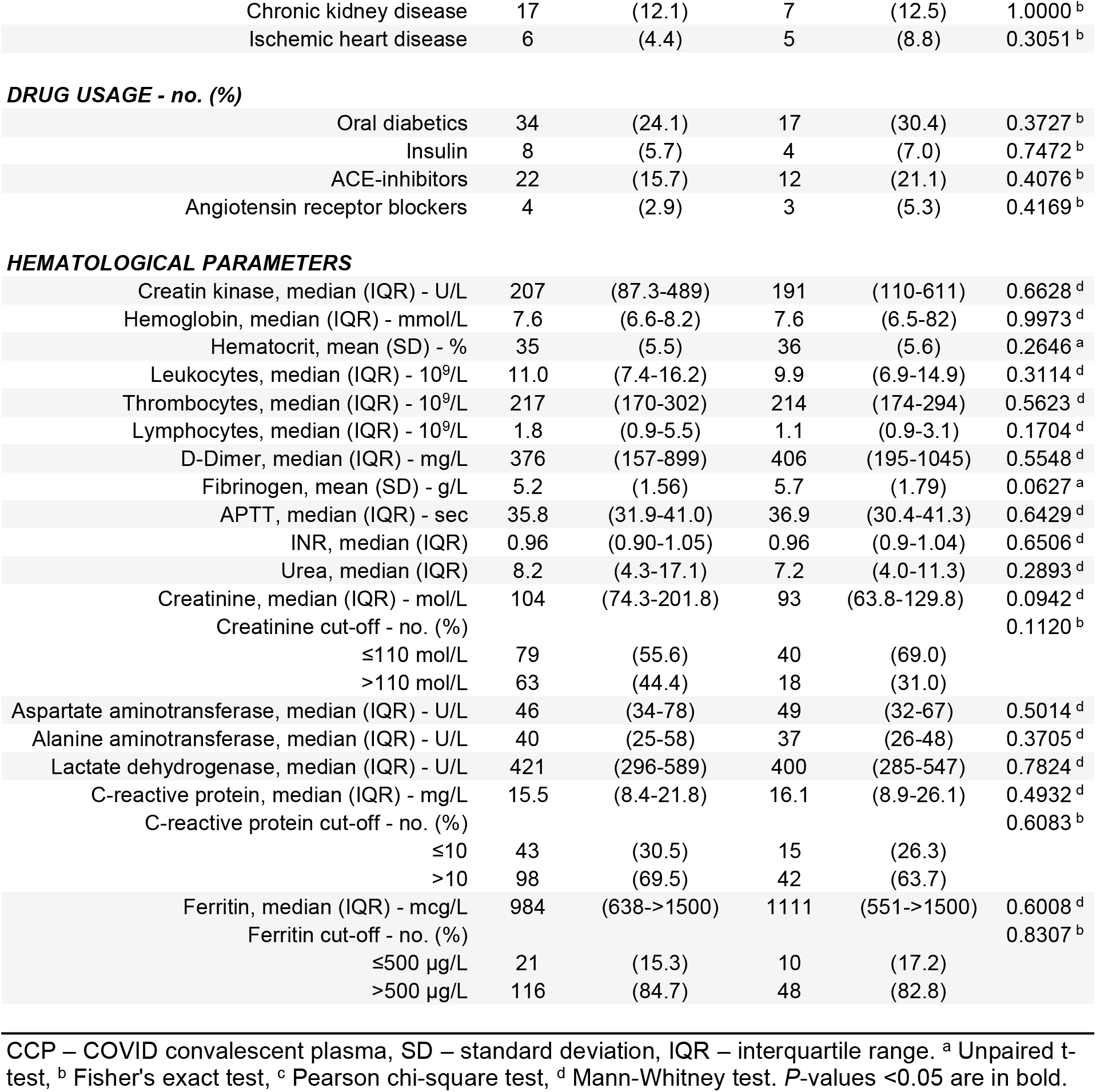
Demographic and clinical characteristics of the patients at baseline.

| Variable | Control<br>(N=142) |  | CCP<br>(N=58) |  | P-value |
| --- | --- | --- | --- | --- | --- |
| DEMOGRAPHICS |  |  |  |  |  |
| Age, mean (SD) - yr | 52 | (14.0) | 50 | (13.1) | 0.3773 <sup>a</sup> |
| Age cut-off - no. (%) |  |  |  |  | 0.6413 <sup>b</sup> |
| 18-50 yr | 63 | (44.4) | 28 | (48.3) |  |
| >50 yr | 79 | (55.6) | 30 | (51.7) |  |
| Male sex - no. (%) | 79 | (55.6) | 31 | (53.4) | 0.8756 <sup>b</sup> |
| Race or ethnic group - no. (%) |  |  |  |  | 0.3655 <sup>c</sup> |
| Javanese | 42 | (29.6) | 20 | (34.5) |  |
| Maroon | 21 | (14.8) | 9 | (15.5) |  |
| Hindustani | 29 | (20.4) | 17 | (29.3) |  |
| Creole | 25 | (17.6) | 9 | (15.5) |  |
| Amerindian | 11 | (7.7) | 1 | (1.7) |  |
| Mix | 13 | (9.2) | 2 | (3.4) |  |
| Other | 1 | (0.7) | 0 | (0) |  |
| BMI, median (IQR) | 28 | (25.7-33.8) | 32 | (26.9-36.3) | **0.0338** <sup>d</sup> |
| BMI cut-off - no (%) |  |  |  |  | **0.0052** <sup>b</sup> |
| ≤30 | 85 | (59.9) | 22 | (37.9) |  |
| >30 | 57 | (40.1) | 36 | (62.1) |  |
| COVID classification severe | 142 | 100% | 58 | 100% | N/A |
| PRESENCE OF SYMPTOMS - no. (%) |  |  |  |  |  |
| Fever | 113 | (79.6) | 47 | (82.5) | 0.6976 <sup>b</sup> |
| Shortness of breath | 129 | (90.8) | 53 | (93.0) | 0.7825 <sup>b</sup> |
| Cough | 102 | (71.8) | 40 | (70.2) | 1.0000 <sup>b</sup> |
| Sputum | 24 | (17.0) | 9 | (15.8) | 1.0000 <sup>b</sup> |
| Loss of taste | 29 | (20.6) | 11 | (19.3) | 1.0000 <sup>b</sup> |
| Loss of smell | 20 | (14.2) | 9 | (15.8) | 0.8252 <sup>b</sup> |
| Itching eyes | 3 | (2.1) | 2 | (3.5) | 0.6269 <sup>b</sup> |
| Red eyes | 2 | (1.4) | 1 | (1.8) | 1.0000 <sup>b</sup> |
| Diarrhea | 30 | (21.3) | 14 | (24.6) | 0.7060 <sup>b</sup> |
| Fatigue | 46 | (32.6) | 27 | (47.4) | 0.0729 <sup>b</sup> |
| Muscle pain | 46 | (32.6) | 30 | (52.6) | **0.0102** <sup>b</sup> |
| Malaise | 23 | (23.7) | 3 | (18.8) | 1.0000 <sup>b</sup> |
| Acute tubular necrosis | 24 | (17.1) | 10 | (18.2) | 0.8369 <sup>b</sup> |
| Vomiting | 19 | (13.6) | 7 | (12.3) | 1.0000 <sup>b</sup> |
| Sore throat | 12 | (18.6) | 5 | (10.6) | 1.0000 <sup>b</sup> |
| Rhabdomyolysis | 2 | (1.4) | 2 | (3.5) | 0.5808 <sup>b</sup> |
| Arrhythmias | 6 | (4.3) | 3 | (5.3) | 0.7198 <sup>b</sup> |
| TIME INTERVALS AT BASELINE |  |  |  |  |  |
| Days hospital until ICU, median (IQR) | 1.0 | (0-3) | 1.0 | (0-5) | 0.2671 <sup>d</sup> |
| Days symptom onset until ICU, median (IQR) | 6.0 | (3-8) | 6.0 | (4-8) | 0.8065 <sup>d</sup> |
| Days illness until treatment start, median (IQR) | 4.0 | (2-7) | 5.0 | (3-8) | 0.2735 <sup>d</sup> |
| Days illness until treatment start cut-off - no. (%) |  |  |  |  | 0.1683 <sup>b</sup> |
| ≤7 days | 116 | (82.9) | 42 | (73.7) |  |
| >7 days | 24 | (17.1) | 15 | (26.3) |  |
| CO-MORBIDITIES - no. (%) |  |  |  |  |  |
| Diabetes mellitus | 38 | (27.0) | 19 | (33.3) | 0.3894 <sup>b</sup> |
| Hypertension | 59 | (42.1) | 26 | (45.6) | 0.7513 <sup>b</sup> |
| Stroke | 5 | (3.6) | 1 | (1.8) | 0.6746 <sup>b</sup> |
| Chronic kidney disease | 17 | (12.1) | 7 | (12.5) | 1.0000<sup>b</sup> |
| Ischemic heart disease | 6 | (4.4) | 5 | (8.8) | 0.3051<sup>b</sup> |
| DRUG USAGE - no. (%) |  |  |  |  |  |
| Oral diabetics | 34 | (24.1) | 17 | (30.4) | 0.3727<sup>b</sup> |
| Insulin | 8 | (5.7) | 4 | (7.0) | 0.7472<sup>b</sup> |
| ACE-inhibitors | 22 | (15.7) | 12 | (21.1) | 0.4076<sup>b</sup> |
| Angiotensin receptor blockers | 4 | (2.9) | 3 | (5.3) | 0.4169<sup>b</sup> |
| HEMATOLOGICAL PARAMETERS |  |  |  |  |  |
| Creatin kinase, median (IQR) - U/L | 207 | (87.3-489) | 191 | (110-611) | 0.6628<sup>d</sup> |
| Hemoglobin, median (IQR) - mmol/L | 7.6 | (6.6-8.2) | 7.6 | (6.5-82) | 0.9973<sup>d</sup> |
| Hematocrit, mean (SD) - % | 35 | (5.5) | 36 | (5.6) | 0.2646<sup>a</sup> |
| Leukocytes, median (IQR) - 10<sup>9</sup>/L | 11.0 | (7.4-16.2) | 9.9 | (6.9-14.9) | 0.3114<sup>d</sup> |
| Thrombocytes, median (IQR) - 10<sup>9</sup>/L | 217 | (170-302) | 214 | (174-294) | 0.5623<sup>d</sup> |
| Lymphocytes, median (IQR) - 10<sup>9</sup>/L | 1.8 | (0.9-5.5) | 1.1 | (0.9-3.1) | 0.1704<sup>d</sup> |
| D-Dimer, median (IQR) - mg/L | 376 | (157-899) | 406 | (195-1045) | 0.5548<sup>d</sup> |
| Fibrinogen, mean (SD) - g/L | 5.2 | (1.56) | 5.7 | (1.79) | 0.0627<sup>a</sup> |
| APTT, median (IQR) - sec | 35.8 | (31.9-41.0) | 36.9 | (30.4-41.3) | 0.6429<sup>d</sup> |
| INR, median (IQR) | 0.96 | (0.90-1.05) | 0.96 | (0.9-1.04) | 0.6506<sup>d</sup> |
| Urea, median (IQR) | 8.2 | (4.3-17.1) | 7.2 | (4.0-11.3) | 0.2893<sup>d</sup> |
| Creatinine, median (IQR) - mol/L | 104 | (74.3-201.8) | 93 | (63.8-129.8) | 0.0942<sup>d</sup> |
| Creatinine cut-off - no. (%) |  |  |  |  | 0.1120<sup>b</sup> |
| ≤110 mol/L | 79 | (55.6) | 40 | (69.0) |  |
| >110 mol/L | 63 | (44.4) | 18 | (31.0) |  |
| Aspartate aminotransferase, median (IQR) - U/L | 46 | (34-78) | 49 | (32-67) | 0.5014<sup>d</sup> |
| Alanine aminotransferase, median (IQR) - U/L | 40 | (25-58) | 37 | (26-48) | 0.3705<sup>d</sup> |
| Lactate dehydrogenase, median (IQR) - U/L | 421 | (296-589) | 400 | (285-547) | 0.7824<sup>d</sup> |
| C-reactive protein, median (IQR) - mg/L | 15.5 | (8.4-21.8) | 16.1 | (8.9-26.1) | 0.4932<sup>d</sup> |
| C-reactive protein cut-off - no. (%) |  |  |  |  | 0.6083<sup>b</sup> |
| ≤10 | 43 | (30.5) | 15 | (26.3) |  |
| >10 | 98 | (69.5) | 42 | (63.7) |  |
| Ferritin, median (IQR) - mcg/L | 984 | (638->1500) | 1111 | (551->1500) | 0.6008<sup>d</sup> |
| Ferritin cut-off - no. (%) |  |  |  |  | 0.8307<sup>b</sup> |
| ≤500 µg/L | 21 | (15.3) | 10 | (17.2) |  |
| >500 µg/L | 116 | (84.7) | 48 | (82.8) |  |

CCP – COVID convalescent plasma, SD – standard deviation, IQR – interquartile range. <sup>a</sup> Unpaired t-test, <sup>b</sup> Fisher's exact test, <sup>c</sup> Pearson chi-square test, <sup>d</sup> Mann-Whitney test. *P*-values <0.05 are in bold.

Convalescent plasma donor recruitment, plasma preparation and data collection were performed as described previously (17).

### Statistical analysis

Unless stated otherwise, all analyses are performed on the complete dataset. General descriptive statistics were assessed using IBM SPSS Statistics for Windows (version 29.0.0.0). Differences between the CCP and SOC treatment groups were analyzed with Pearson chi-Square or Fisher exact tests where suitable for categorical variables, T tests for parametric continuous variables and Mann Whitney tests for non-parametric continuous variables. The differences in outcome measures were analyzed using univariate and multivariate cox proportional hazard analyses. The mortality risk was assessed using Kaplan Meier survival analysis and hazard ratios calculated with a cox proportional hazard model. CXR score and P/F ratio were summarized by representing the median and spread in a boxplot and differences between the CCP and SOC treatment groups analyzed with Mann Whitney tests. A *P*-value of 0.05 was considered to represent significant differences. Graphs of study data were generated using GraphPad Prism (version 9.4.1).

## Results

### Participants

A total of 200 patients were enrolled in the study of which 58 (29%) received CCP. The mean age of the patients was 51.6±13.7 years and 110 patients (55%) were male. The demographic, clinical and hematological characteristics are described in **Table 1**. The intervention (CCP) and control (SOC) treatment groups were not significantly different for the vast majority of baseline parameters (demographics, presence of symptoms, co-morbidities, medication, and hematological parameters, see **Table 1**). The CCP and SOC treatment groups did show significant differences in BMI (median BMI 32 (27-36) versus 28 (26-34), respectively, Mann Whitney *P* = 0.0338) and muscle pain (53% versus 33%, respectively, Fisher’s exact *P* = 0.0102). The CCP and SOC treatment groups did not show significant differences for baseline hospital parameters such as days of symptoms or days in hospital until ICU admission, and days of illness until start of treatment.

### Primary outcome: mortality

CCP treatment was associated with significantly fewer deaths: 12 of 58 (21%) patients in the CCP treatment group died versus 56 of 142 (39%) patients in the SOC group, Fisher’s exact *P* = 0.0133 (see **Table 2**). Days spent in the hospital or ICU were significantly higher in the CCP treatment group. These results confirm the protective effect previously reported for the pre-planned interim analysis of this trial (17).

**Table 2.**
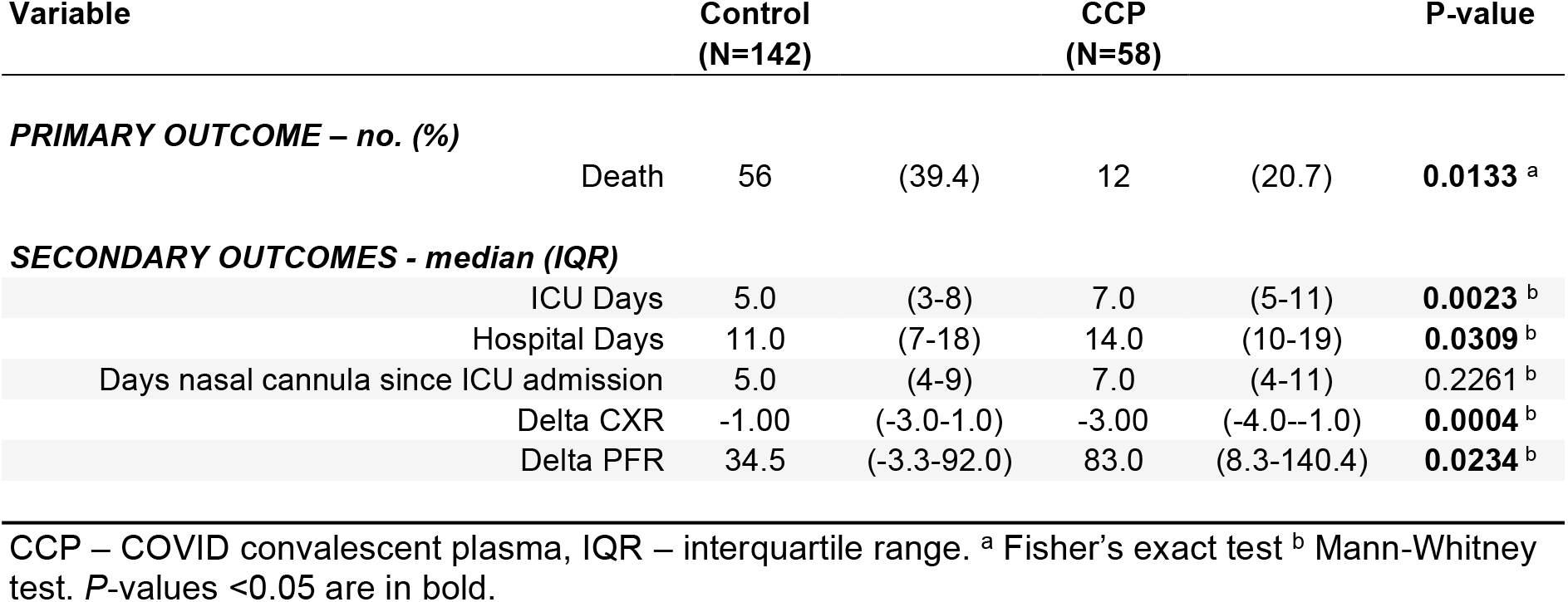
Disease severity in hospitalized patients.

| Variable | Control<br>(N=142) |  | CCP<br>(N=58) |  | P-value |
| --- | --- | --- | --- | --- | --- |
| PRIMARY OUTCOME – no. (%) |  |  |  |  |  |
| Death | 56 | (39.4) | 12 | (20.7) | 0.0133 <sup>a</sup> |
| SECONDARY OUTCOMES - median (IQR) |  |  |  |  |  |
| ICU Days | 5.0 | (3-8) | 7.0 | (5-11) | 0.0023 <sup>b</sup> |
| Hospital Days | 11.0 | (7-18) | 14.0 | (10-19) | 0.0309 <sup>b</sup> |
| Days nasal cannula since ICU admission | 5.0 | (4-9) | 7.0 | (4-11) | 0.2261 <sup>b</sup> |
| Delta CXR | -1.00 | (-3.0-1.0) | -3.00 | (-4.0--1.0) | 0.0004 <sup>b</sup> |
| Delta PFR | 34.5 | (-3.3-92.0) | 83.0 | (8.3-140.4) | 0.0234 <sup>b</sup> |

CCP – COVID convalescent plasma, IQR – interquartile range. <sup>a</sup> Fisher's exact test <sup>b</sup> Mann-Whitney test. *P*-values <0.05 are in bold.

### Univariate hazard analyses

In addition to differences identified between the groups at baseline (BMI and muscle pain), other factors that may affect in-hospital mortality have been reported, such as age, sex, ethnicity, diabetes mellitus status, timing of CCP intervention, and levels of creatinine, C-reactive protein, ferritin, and fibrinogen. The effects these factors may have on the primary outcome were evaluated using univariate hazard analyses (**Table 3** and **Figure 2**). Possible confounders BMI and muscle pain (see **Table 1**) did not affect hazard ratios and were not included in the subsequent multivariate analyses. Using number of ICU days as marker, in addition to treatment type (CCP versus SOC), age and creatinine levels significantly affected hazard ratios (see **Table 3**). Using number of hospital days as marker, in addition to treatment type, also age, diabetes mellitus status, timing of treatment start and creatinine levels significantly affected hazard ratios (see **Table 3**). Next, these factors were included in the multivariate analyses.

**Table 3.** Univariate hazard ratios for death.

| Factor | ICU Days <sup>b</sup> |  |  | Hospital Days <sup>c</sup> |  |  |
| --- | --- | --- | --- | --- | --- | --- |
|  | HR (exp(B)) | 95%CI | P-value | HR (exp(B)) | 95%CI | P-value |
| Age (cut-off at 50) | 2.110 | 1.216-3.663 | **0.008** | 2.256 | 1.300-3.913 | **0.004** |
| BMI (cut-off at 30) | 0.871 | 0.537-1.413 | 0.576 | 0.875 | 0.540-1.417 | 0.587 |
| Sex (male-female) | 0.818 | 0.503-1.331 | 0.418 | 0.797 | 0.490-1.296 | 0.360 |
| Ethnicity (1-7) <sup>a</sup> | 1.100 | 0.941-1.284 | 0.231 | 1.001 | 0.862-1.162 | 0.994 |
| Muscle pain (no or yes) | 0.817 | 0.487-1.372 | 0.445 | 0.726 | 0.433-1.219 | 0.226 |
| Diabetes Mellitus (no or yes) | 1.465 | 0.901-2.383 | 0.124 | 1.687 | 1.039-2.737 | **0.034** |
| Treatment start (cut-off at 7) | 0.579 | 0.294-1.141 | 0.114 | 0.467 | 0.236-0.924 | **0.029** |
| Therapy type (SOC or CCP) | 0.354 | 0.188-0.663 | **0.001** | 0.464 | 0.248-0.868 | **0.016** |
| Creatinine (cut-off at 110) | 4.945 | 2.884-8.479 | **<0.001** | 4.506 | 2.658-7.640 | **<0.001** |
| CRP (cut-off at 10) | 1.007 | 0.557-1.821 | 0.981 | 1.494 | 0.824-2.708 | 0.186 |
| Ferritin (cut-off at 500) | 1.372 | 0.591-3.183 | 0.462 | 1.980 | 0.855-4.589 | 0.111 |
| Fibrinogen (continuous) | 0.973 | 0.853-1.111 | 0.690 | 0.968 | 0.840-1.114 | 0.649 |

ICU – intensive care unit, HR – hazard ratio, SOC – standard of care, CCP – COVID convalescent plasma. <sup>a</sup> Ethnicity: 1 – Javanese, 2 – Maroon, 3 – Hindustani, 4 – Creole, 5 – Amerindian, 6 – Mix, 7 – Other. <sup>b</sup> 28-day follow up. <sup>c</sup> 35-day follow up. *P*-values <0.05 are in bold.

**Figure 2.**
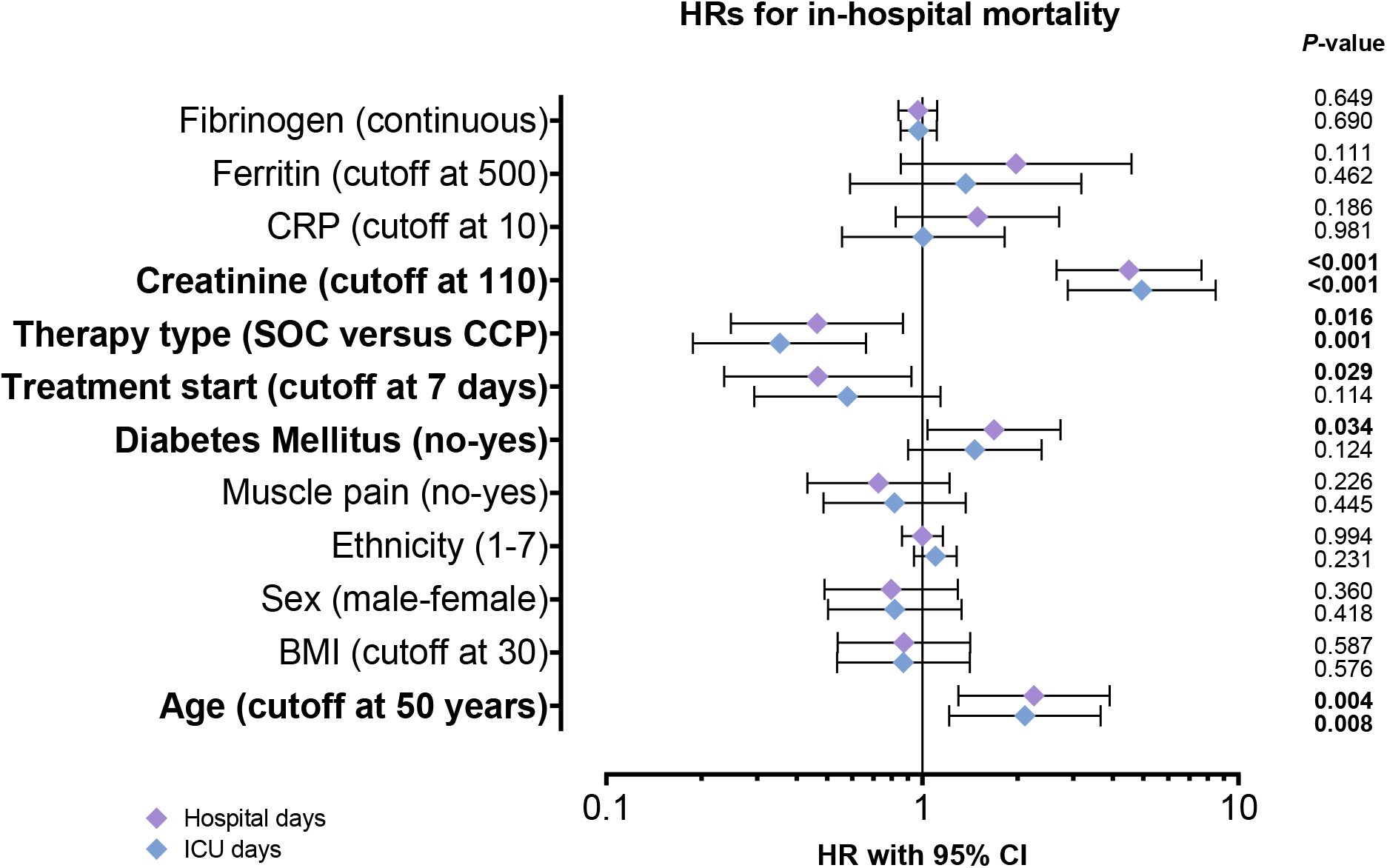
Identification of covariates to be included in multivariate analyses. Univariate cox proportional analyses of hospital days (purple diamonds) and ICU days (blue diamonds) was performed for factors that may affect in-hospital mortality. Graph depicts hazard ratios with 95% confidence intervals. Log-rank *P*-values are shown. Bolded variables are significantly associated with death and included in the multivariate cox proportional hazard analyses.

### Multivariate hazard analysis

In a covariates-adjusted Cox model using ICU days, convalescent plasma transfusion was significantly associated with improved survival (*P* = 0.001), independent of creatinine levels (*P* < 0.001)(**Table 4 and Figure 3**). Similarly, in the model using hospital days, convalescent plasma transfusion was significantly associated with improved survival (*P* = 0.030), independent of creatinine levels (*P* < 0.001) and timing of treatment start (*P* = 0.015)(**Table 4 and Figure 3**).

**Table 4.**
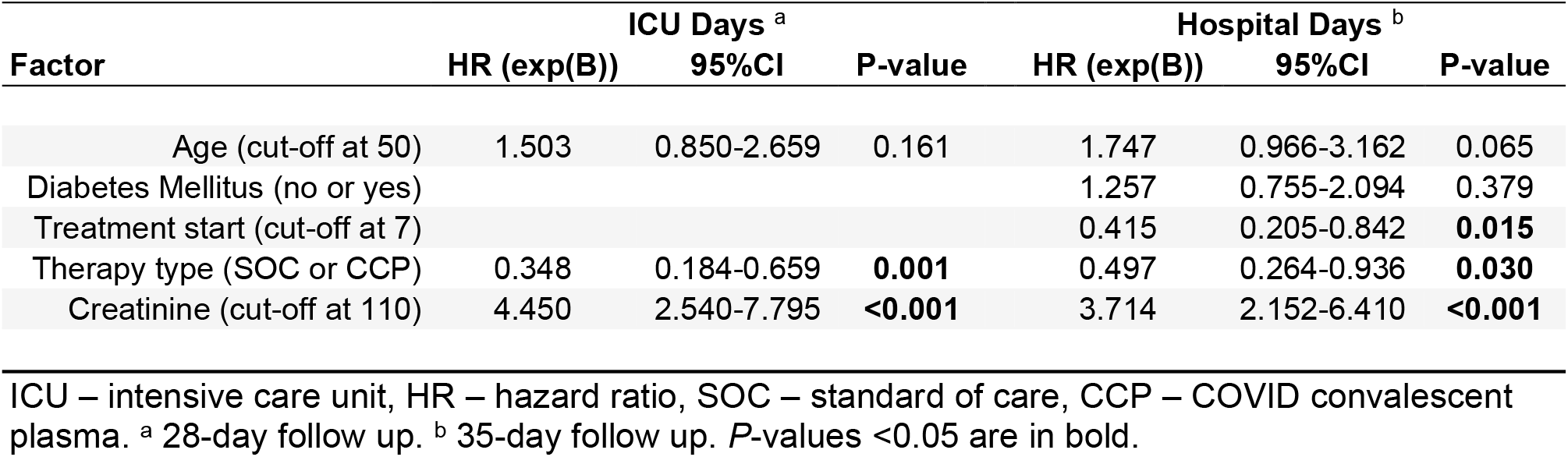
Multivariate hazard ratios for death.

| Factor | ICU Days <sup>a</sup> |  |  | Hospital Days <sup>b</sup> |  |  |
| --- | --- | --- | --- | --- | --- | --- |
|  | HR (exp(B)) | 95%CI | P-value | HR (exp(B)) | 95%CI | P-value |
| Age (cut-off at 50) | 1.503 | 0.850-2.659 | 0.161 | 1.747 | 0.966-3.162 | 0.065 |
| Diabetes Mellitus (no or yes) |  |  |  | 1.257 | 0.755-2.094 | 0.379 |
| Treatment start (cut-off at 7) |  |  |  | 0.415 | 0.205-0.842 | **0.015** |
| Therapy type (SOC or CCP) | 0.348 | 0.184-0.659 | **0.001** | 0.497 | 0.264-0.936 | **0.030** |
| Creatinine (cut-off at 110) | 4.450 | 2.540-7.795 | **<0.001** | 3.714 | 2.152-6.410 | **<0.001** |

ICU – intensive care unit, HR – hazard ratio, SOC – standard of care, CCP – COVID convalescent plasma. <sup>a</sup> 28-day follow up. <sup>b</sup> 35-day follow up. *P*-values <0.05 are in bold.

**Figure 3.**
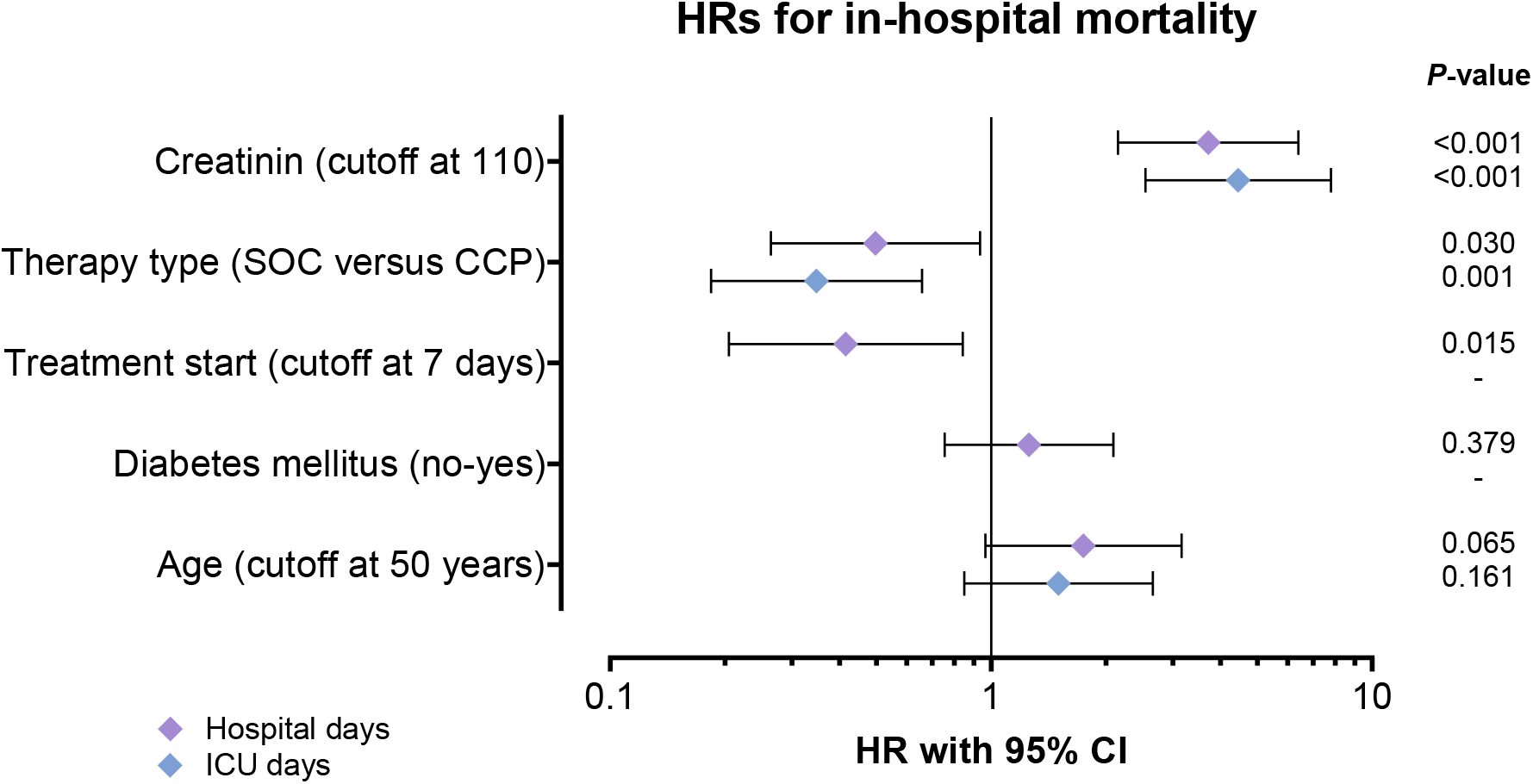
Multivariate analyses of covariates for in-hospital mortality. A multivariate cox proportional hazard analysis was performed with the univariately significant variables for hospital days (purple diamonds) and ICU days (blue diamonds). Graph depicts hazard ratios with 95% confidence intervals. Log-rank *P*-values are shown.

### Survival analyses

As of the end of the study, 21% of the CCP treatment group and 39% of the SOC treatment group had died (**Table 2**). The median ICU follow-up time was 7 days (range, 1-27 days) for the CCP treatment group and 5 days (range, 0-50 days) for the SOC treatment group. Median hospital follow-up time was 14 days (range, 4-37 days) and 11 days (range, 1-62 days), respectively. Using either ICU days or hospital days survival probability was significantly greater in the CCP than in the SOC treatment group (**Figure 4**). Without covariate adjustment, the survival benefit using ICU days had a cox hazard ration of 0.354 (0.188-0.663) with a log-rank (Mantel-Cox) *P*-value of 0.0005 (**Figure 4A**). Similarly, using hospital days the survival benefit had a cox hazard ratio of 0.464 (0.248-0.868) with a log-rank (Mantel-Cox) *P*-value of 0.0127 (**Figure 4B**).

**Figure 4.**
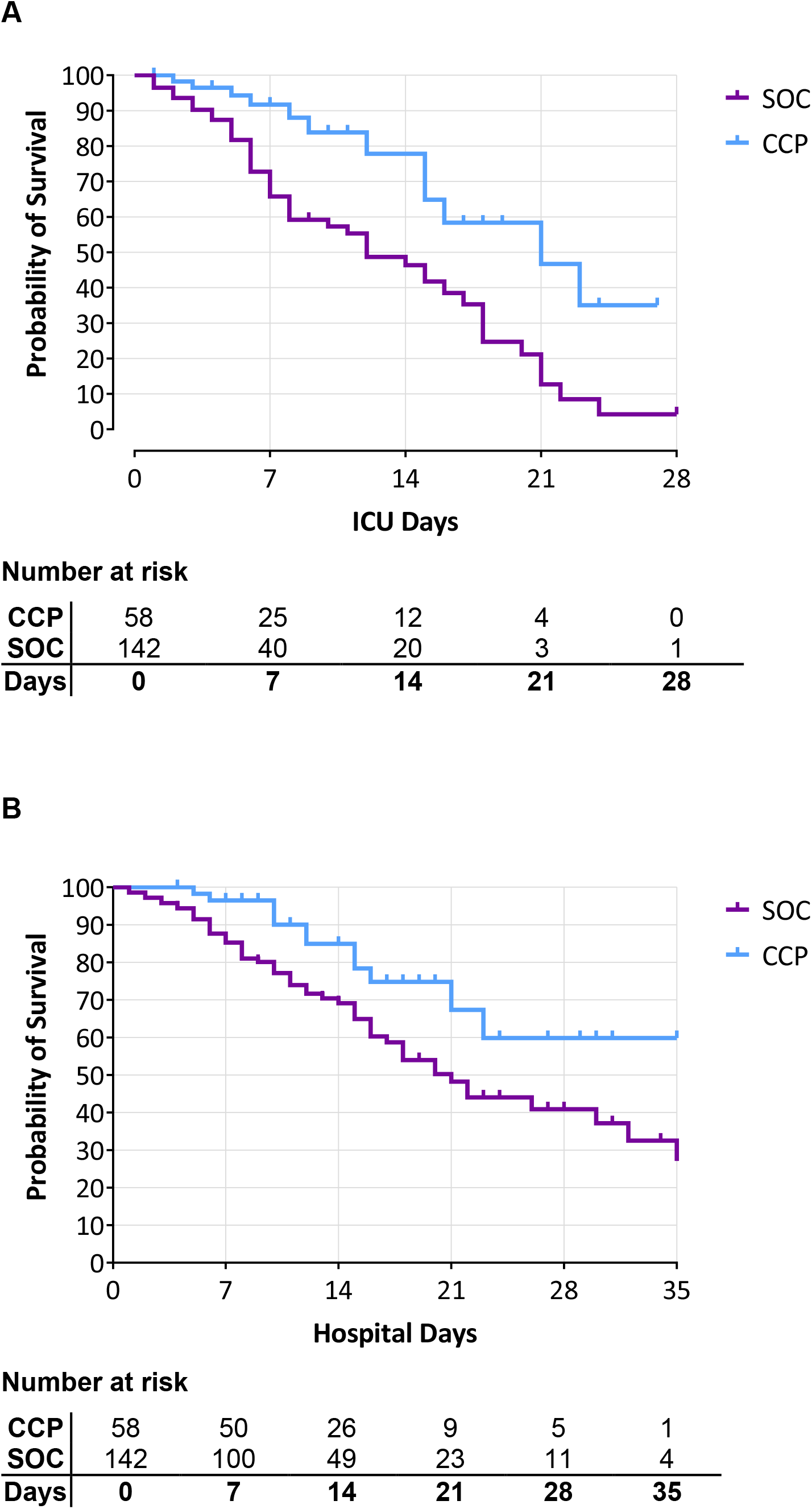
Survival probability during hospital admission for CCP and SOC treatment groups. (**A**) Kaplan Meier curve of ICU days for CCP and SOC treatment groups. Right-censoring took place when a person was dismissed from the ICU before the last measured timepoint, death was counted as an event. Log Rank (Mantel-Cox) *P*-value < 0.001. The longest ICU admission duration in the CCP group was 27 days, while it was 50 days in the SOC group. (**B**) Kaplan Meier curve of hospital days for CCP and SOC treatment groups. Right-censoring took place when a person was dismissed from hospital before the last measured timepoint, death was counted as an event. Log Rank (Mantel-Cox) *P*-value = 0.013. The longest hospital admission duration in the CCP group was 37 days, while it was 62 days in the SOC group.

### Secondary outcomes: CXR score and P/F ratio

Chest radiographic findings (CXR score): To identify early changes in clinical response, the CXR score was determined for each patient upon ICU admission (day 0) and 48 hours after the treatment initiation (day 2). The difference in CXR score between day 0 and day 2 was calculated (delta CXR) and compared between the two groups (**Figure 5A**). The delta CXR after CCP treatment (median −3 points, IQR −4 to −1) was significantly greater than in the SOC group (median −1 point, IQR −3 to 1), Mann-Whitney *P* = 0.0004 (**Table 2**).

**Figure 5.**
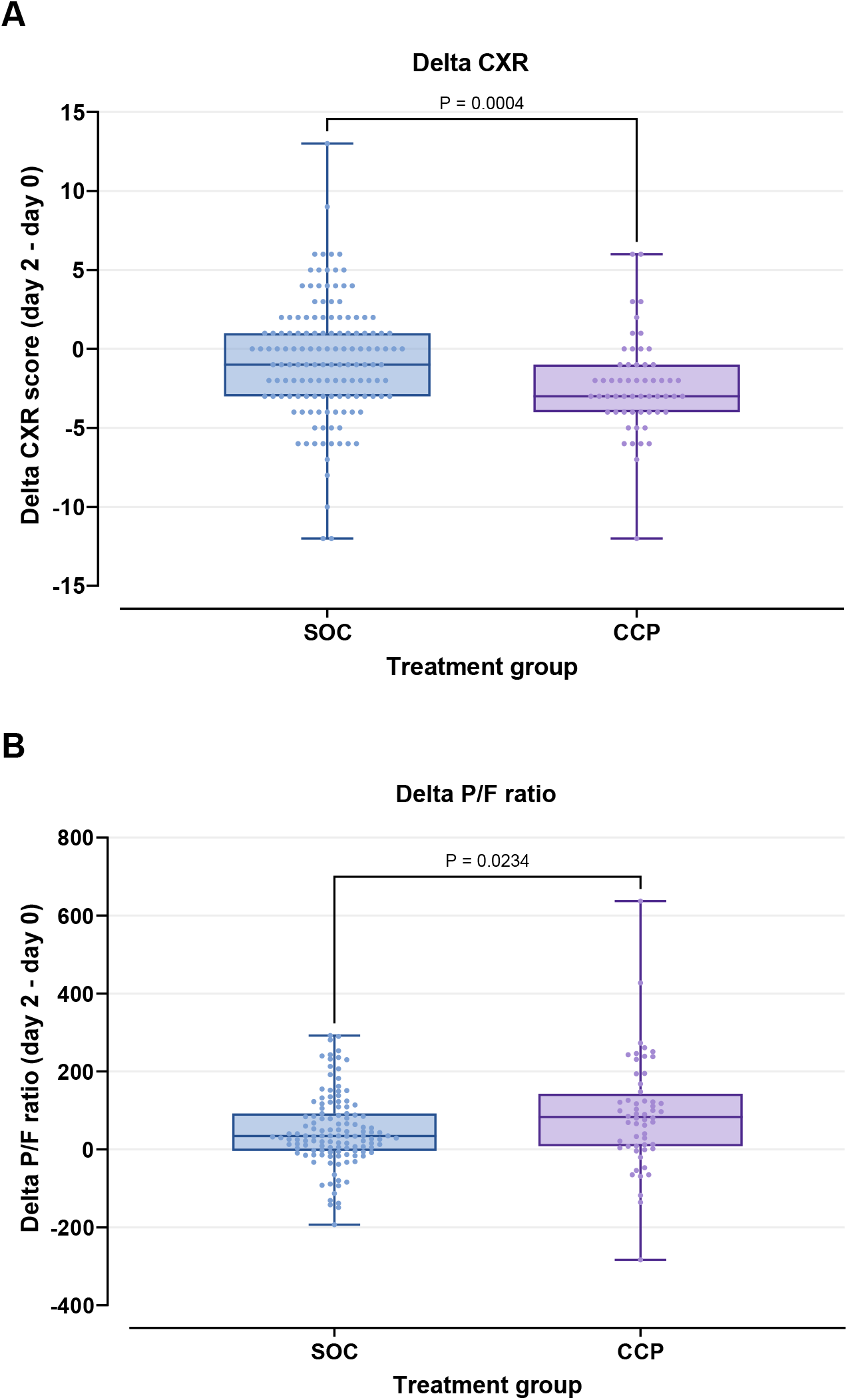
CXR score and P/F ratio improvement in CCP and SOC treatment groups. The delta CXR and delta PFR were each calculated by subtracting the respective scores of day 0 from the score on day 2. Graphs show minimum, median, interquartile range and maximum for each group. **(A)** The CCP group showed significantly greater improvement in CXR score than the SOC group (Mann-Whitney test). **(B)** The CCP group showed significantly greater improvement in P/F ratio than the SOC group (Mann-Whitney test).

Pulmonary oxygen exchange capacity (P/F ratio): The P/F ratio was also determined for each patient upon ICU admission and 48 hours after treatment initiation. The difference in P/F ratio between day 0 and day 2 was calculated (delta PFR) and compared between the two groups (**Figure 5B**). The delta PFR after CCP treatment, (median 83, IQR 8 to 140) was significantly greater than in the SOC group (median 35, IQR −3 to 92), Mann-Whitney *P* = 0.0234 (**Table 2**).

## Discussion

This study compares mortality and other endpoints between ICU COVID-19 patients treated with convalescent plasma plus standard of care (CCP), and a control group of patients hospitalized in the same medical ICU facility treated with standard of care alone (SOC) in a LMIC setting. It demonstrates a significant survival improvement in CCP recipients (HR 0.35; 95% CI, 0.19–0.66; *P* = 0.001).

Early in the pandemic, Duan and colleagues (20) reported the potential beneficial effect of CCP showing improvements in respiratory and chest X ray parameters without serious side effects. Improved ICU survival was also reported in this early phase of the pandemic (21). Limited treatment options and the absence of vaccines at that time made CCP an attractive alternative. Several ICU outcome studies followed with similar results: e.g. Tworek and colleagues (22) reported >50% reduction in mortality in the CCP group for a propensity-score matched case control study in Poland, an effect size similar to that of age and timing of treatment initiation. Equivalent mortality rates were reported for patients in Wuhan (23), indicating comparable ICU treatment levels between LMIC versus high income countries (HIC) with limited resources and capacity (24).

Liu and colleagues (7) showed that CCP was beneficial in patients with severe or life-threatening COVID-19 disease. These CCP patients experienced a significant improvement of clinical symptoms - such as a reduction of oxygen requirement ratio - as well as survival over SOC patients (HR 0.34; 95% CI, 0.13–0.89; *P* = 0.027). But other studies showed limited or even negative results with CCP. Altuntas and colleagues (25) matched a large retrospective cohort of >1600 ICU patients with or without CPP treatment, showing an improvement of clinical symptoms but only a modest effect on mortality (24.7 % in the CP group, and 27.7 % in the control group). In one of the rare randomized studies, Simonovich and colleagues (12) could not detect significant differences in mortality rate between CCP treated patients and those who received placebo. However, mortality in their SOC and CCP treatment groups was 11.4% and 11.0%, respectively. In addition, most patients were enrolled from the hospital ward (73.3% and 65.8%, respectively) instead of including ICU admitted patients only. This may well explain the low mortality compared to our patient cohort, indicating a possible hurdle to detecting the benefit of CCP treatment in their study population.

The failure of CCP treatment to improve disease outcome in modestly ill patients in the multicenter randomized trial reported by Agarwal and colleagues (13) severely affected acceptance of CCP treatment and led to its ban in India. The CCP treatment failure was correlated with the use of low-titer CCP, since confirmed in multiple studies. Moreover, studies from the USA showed that high-titer CCP and early treatment initiation resulted in improved mortality reduction (26, 27)(28). However, also in the USA, the use of CCP as an emergency drug was restricted and limited to immunocompromised patients only (29). Despite the conflicting results summarized above, promising results from our initial study (17), led us to continue the evaluation of CPP treatment in ICU patients. This study indeed confirmed the efficacy of CCP treatment in our setting.

The efficacy of CCP treatment is also affected by the CCP source donors and its methods of collection/production - which are not standardized and therefore do not guarantee its anti-viral contents (30, 31). This might be one of the reasons for the strong efficacy of the CCP used in our setting: CCP was donated by donors early after their recovery and collected using an innovative blood separator, HemoClear, which allows higher particle content of the plasma (18). Conventional plasmapheresis produces plasma particles with an upper limit of 0.4μm. In contrast, the HemoClear method upper limit is 2.3μm allowing also high molecular proteins and 30% circulating platelets to be gathered into the plasma. High Molecular weight proteins such as IgM and IgG complexes are between the range of 0.2-0.5μm and may be inactivated or damaged by a conventional CCP method (32–34).

Platelets in the CCP may have important immune modulatory and protective effects. After the collection, freezing and thawing of the CCP, platelets are a component of plasma produced using the HemoClear device. Plasma also contains a variety of extracellular vesicles - cell-released, membranous structures – called exosomes, microvesicles, or microparticles, among others (35). Studies are ongoing to explore extracellular vesicles as a disease modifying biotherapy (36) and may have contributed to the efficacy of the HemoClear-produced CCP, as reflected in both the early signs of recovery - measured by CXR and P/F ratio - and improved survival rate.

Another advantage of the HemoClear device is the reduced cost compared to conventional plasmapheresis machines. While the costs for patient identification, contact, and antibody testing are the same for both methods, initial investment (around $25,000 per machine) and technical staff requirements are higher with conventional plasmapheresis (37). Moreover, due to the 60-90 minute processing time each plasmapheresis machine is limited to four collections daily, thus limiting sites in their ability to adjust CCP production to local need. In the USA, federal contracts worth $646 million were paid to U.S. blood centers to collect 500,000 units of covid convalescent plasma, meaning a unit costs of $1,300 for the USA government (38). Due to the added complexity of CCP, blood centers have been reimbursed $600 to $800 per unit of CCP (38). In contrast, the HemoClear method costs were well below $300 per unit of CCP.

The availability of CCP was not only a problem in LMICs but also in the western world. In the nationwide USA CCP program, more than 30% of the indications was not met due to lack of CCP availability. In addition, CCP programs interfered with the conventional blood donation programs since CCP donors were not available for whole blood donation for 3 months afterwards (39).

Early treatment (within 7 days) after illness onset is paramount in order to treat patients effectively, and this was confirmed in our setting. In order to prevent ICU admission, CCP treatment could even be initiated before ICU admission. Indeed, Libster and colleagues (40) showed a 48% reduction of hospital admission when CCP was used as treatment within the time span of 3 days after becoming ill at home. With bedside production of CCP in primary health care facilities in local communities, not limited by the upfront costs and technological barriers of conventional plasmapheresis, CCP produced by the HemoClear method could be a first line of defense against new virus mutants that are not responsive to existing vaccines, not only in the HICs but also in LMICs (41–43).

The initial lack of prophylactic or therapeutic treatment options combined with limited ICU or even hospital capacity, resulted in decisions to lock down society to reduce viral spread at the start of the pandemic. Decisions not to implement lock downs, e.g. in Brazil and Tanzania, resulted in excessive deaths as well as breakdown of the healthcare system (44–46). Even in these extreme situations, CCP treatment may help reduce healthcare burden and disease mortality.

Although this is an exploratory study, it clearly shows the benefit of using the HemoClear-produced CCP in ICU patients in the Suriname LMIC setting. Additional studies can further substantiate our findings and their applicability to both LMIC and HIC and the use of CCP to combat new viral pandemics.

## Data Availability

All data produced in the present study are available upon reasonable request to the authors.

## Author Contributions

RBI, RBA, SVR, ZCH, DDI and ANI conceived and designed the study. RBI, RBA, NRA, RMA, DBU, EDI, CFU, and ITH conducted the study. RBI, RBA, AWO and ANI analyzed the data and generated figures. All authors reviewed, critically revised, and approved the final version of the manuscript.

## Conflict of Interest

Arno P. Nierich is the inventor of the HemoClear filter and holds stock ownership in HemoClear BV, Dr. Stolteweg 70, 8025 AZ Zwolle, Netherlands. No involvement in actual patient treatment Suriname.

All other authors: no conflict of interest.

## Funding

This study was funded by the Academic Hospital Paramaribo without additional third-party funding.

## Acknowledgements

The authors express their gratitude to the trial participants and to the COVID-19 convalescent plasma donors who generously gave of their time and donated biological specimens. We would like to thank Dion Osemwengi for her support in training on use of the blood filter and contribution to the plasma acquisition protocol.

## Supplementary Materials

Supplementary Video 1: Infographic on the plasma collection method (Link 1: https://www.youtube.com/watch?v=gQGh-PtfBLk&t=20s).

## Notes

### Competing Interest Statement

Arno P. Nierich is the inventor of the HemoClear filter and holds stock ownership in HemoClear BV, Dr. Stolteweg70, 8025 AZ Zwolle, Netherlands. No involvement in actual patient treatment Suriname. All other authors: no conflict of interest.

### Clinical Trial

ISRCTN18304314, ISRCTN49832318

### Author Declarations

Ethical approval is granted by the Suriname Ministry of Health's Ethics Review Board (registration number:IGAP02-482020; ISRCTN18304314).

